# Placental vascular remodeling in pregnant women with COVID-19

**DOI:** 10.1101/2021.07.01.21259860

**Authors:** Sergiy G. Gychka, Iurii L. Kuchyn, Tetyana V. Savchuk, Sofia I. Nikolaienko, Volodymyr M. Zhezhera, Ihor I. Chermak, Yuichiro J. Suzuki

## Abstract

Severe acute respiratory syndrome coronavirus 2 has been causing the pandemic of coronavirus disease 2019 (COVID-19) that has so far resulted in over 180 million infections and nearly 4 million deaths. This respiratory virus uses angiotensin-converting enzyme 2 as a receptor to enter host cells, exhibiting a unique feature that affects various tissues in addition to the lungs. The present study reports that the placental arteries from women who gave birth to live full-term newborns while developing of COVID-19 during pregnancy exhibit severe vascular wall thickening and the occlusion of the vascular lumen. A morphometric analysis of the placental arteries stained with hematoxylin and eosin suggest a 2-fold increase in wall thickness and a 5-fold decrease in the lumen area. Immunohistochemistry with α-smooth muscle actin and Masson’s trichrome staining showed that such placental vascular remodeling in COVID-19 is associated with smooth muscle proliferation and fibrosis. Placental vascular remodeling may represent a mechanism of the clinical problems associated with childbirth in COVID-19 patients.

## Introduction

Coronaviruses are positive-sense, single-stranded RNA viruses that often cause the common cold [1,2]; however, some coronaviruses can promote severe health problems. Currently, severe acute respiratory syndrome coronavirus 2 (SARS-CoV-2) has been causing the pandemic of coronavirus disease 2019 (COVID-19) [3,4]. So far, over 180 million people have been infected with SARS-CoV-2, with nearly 4 million deaths worldwide, causing enormous health, economic, and sociological problems. SARS-CoV-2 enters host cells via the angiotensin-converting enzyme 2 (ACE2) receptor [5,6]. While lung cells are the primary targets of this respiratory virus, causing acute respiratory distress syndrome [7,8], SARS-CoV-2 also affects other ACE2-expressing tissues including those of the cardiovascular system [4,9,10].

COVID-19 is associated with increased neonatal and maternal morbidity and mortality [11-13]. However, the mechanism of the effects of COVID-19 on pregnant women is not understood. Such information should help develop therapeutic strategies to reduce the mortality and morbidity associated with COVID-19.

The present study demonstrates that women who became infected with SARS-CoV-2 and developed COVID-19 symptoms during pregnancy exhibited severe vascular remodeling of the placental arteries as determined by histological examinations. Vascular remodeling is associated with increased smooth muscle cell proliferation and fibrosis. These mechanisms may explain how some neonates are affected by COVID-19.

## Materials and Methods

### Tissue samples

Placental tissues were collected from 113 patients who gave birth to live full-term newborns in Ukraine. Among them, 85 patients were infected with SARS-CoV-2 and developed COVID-19 symptoms at 28 -36 weeks of pregnancy. For comparison purposes, we used placental tissues from 28 women who underwent live childbirth without contracting SARS-CoV-2 as determined by a PCR test. Ages of mothers, weeks of gestation when the child birth occurred, placenta weights, and Apgar scores are summarized in Table 1. Clinical studies were approved by the regional committee for medical research ethics in Kiev, Ukraine and performed under the Declaration of Helsinki of 1975 revised in 2013 or comparable ethical standards. All the patients provided written informed consent.

**Table 1:**
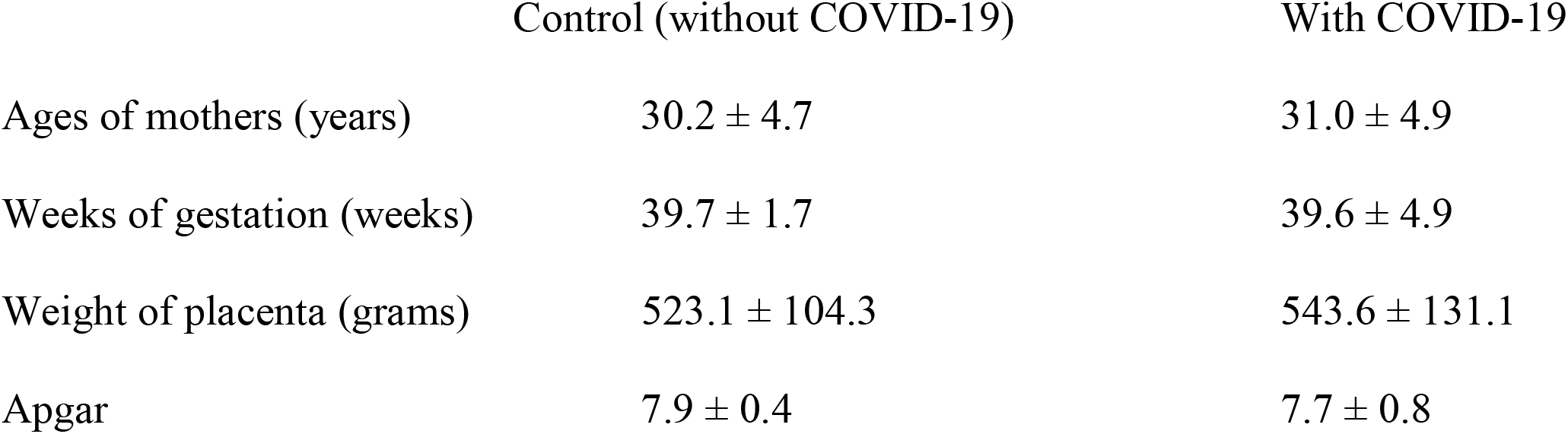
Characteristics of samples used in this study (means ± SD)

|  | Control (without COVID-19) | With COVID-19 |
| --- | --- | --- |
| Ages of mothers (years) | $30.2 \pm 4.7$ | $31.0 \pm 4.9$ |
| Weeks of gestation (weeks) | $39.7 \pm 1.7$ | $39.6 \pm 4.9$ |
| Weight of placenta (grams) | $523.1 \pm 104.3$ | $543.6 \pm 131.1$ |
| Apgar | $7.9 \pm 0.4$ | $7.7 \pm 0.8$ |

### Histological examinations

Tissue samples were fixed overnight in 10% buffered formalin at room temperature. Fixed tissues were embedded in paraffin. From paraffin blocks, 5 μm thick sections were made using a Leica SM 2000 R microtome. The sections were stained with hematoxylin and eosin (H&E) and Masson’s trichrome, and immunohistochemistry using the antibody against α-smooth muscle actin was carried out. Histological specimens were examined using a Leica BX 51 microscope, a Leica MC 190 digital camera, and the Leica LAS software at a magnification of 200x. Morphometric investigations included the assessment of placental arterial wall thickness and the placental arterial lumen index (the ratio of the internal vessel area to the external vessel area) using the ImageJ software.

### Statistical analysis

For the morphometric analysis, IBM SPSS Statistics software version 22.0 was used for the statistical calculations. A Mann-Whitney U test was used to define statistical significance at *p* < 0.05.

## Results

Fig. 1 shows the representative placental arteries of women who gave birth to live full-term newborns. Panels A-C are the H&E images of the placental arteries of represenative three women who gave birth without developing COVID-19. Panels D-F show the H&E images of the placental arteries of representative three women who gave birth but were found to be positive for SARS-CoV-2 and developed symptoms of COVID-19 at 28, 32, and 36 weeks of pregnancy. The placental arteries of women who developed COVID-19 during pregnancy consistently exhibited histological characteristics of vascular wall thickening and vascular lumen narrowing. By contrast, the placental vessels of mothers who did not have COVID-19 during pregnancy showed no occurrences of thickened placental arteries.

**Figure 1:**
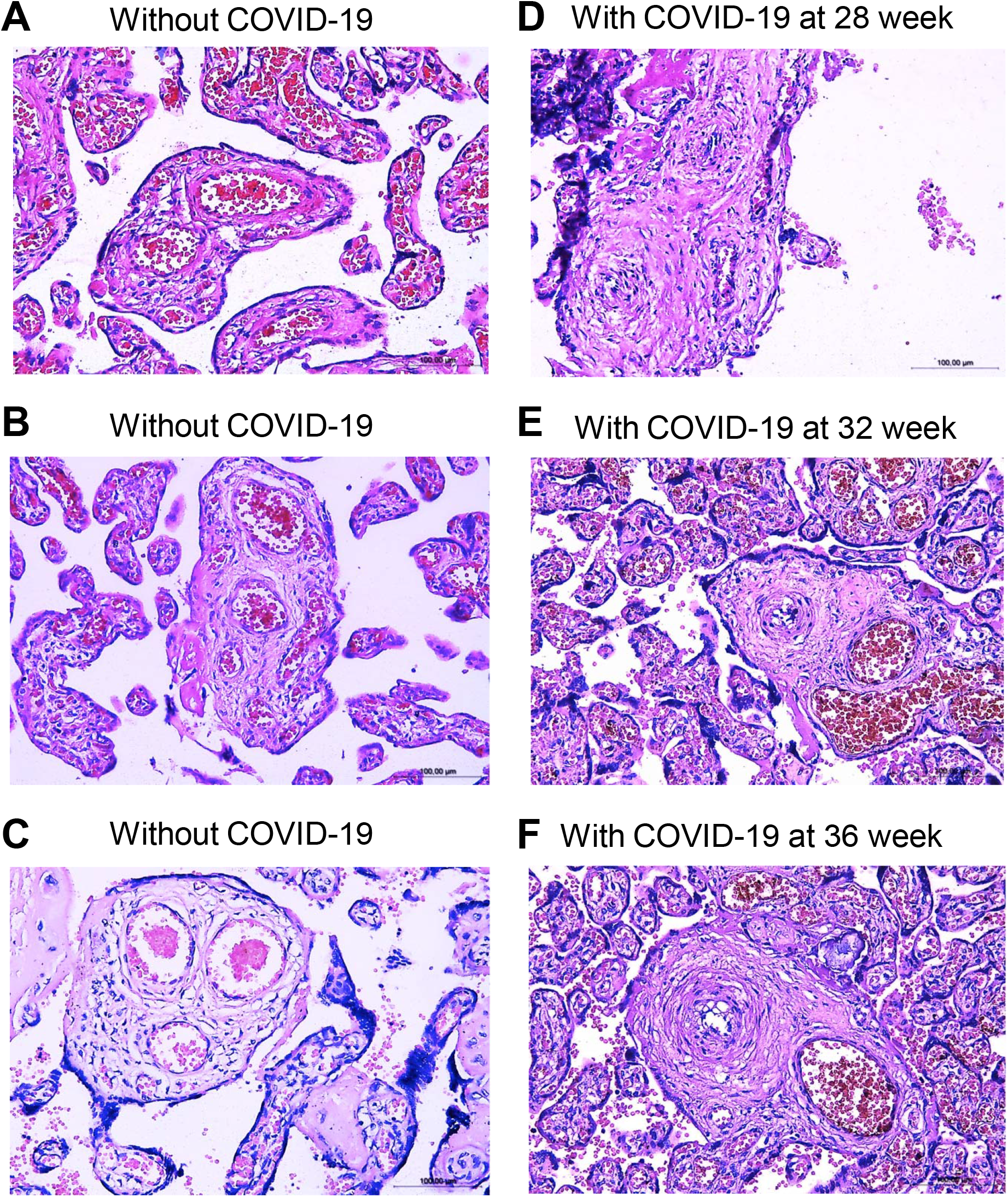
Vascular wall thickening in the placentas of women who had COVID-19 during pregnancy. (A-C) H&E staining images of placental semi-stem villi arteries of representative three women who gave birth without having COVID-19 (40 week of gestation). (D-E) H&E staining images of placental semi-stem villi arteries of representative three women who gave birth (40 weeks of gestation) while contracting COVID-19 at 28, 32 or 36 weeks of gestation. Magnification x200.

The morphometric analysis of placental arterial wall thickness showed that the median value of COVID-19 patients was ∼30 μm (N = 85 placentas), while that of controls was ∼15 μm (N = 28 placentas), indicating that placental arterial walls are twice as thick in COVID-19 patients than in those without COVID-19 (Fig. 2A). These two values are significantly different (*p* < 0.01). The placental arterial lumen area was found to be significantly smaller (5-fold) in COVID-19 patients than in controls (*p* < 0.01, Fig. 2B).

**Figure. 2:**
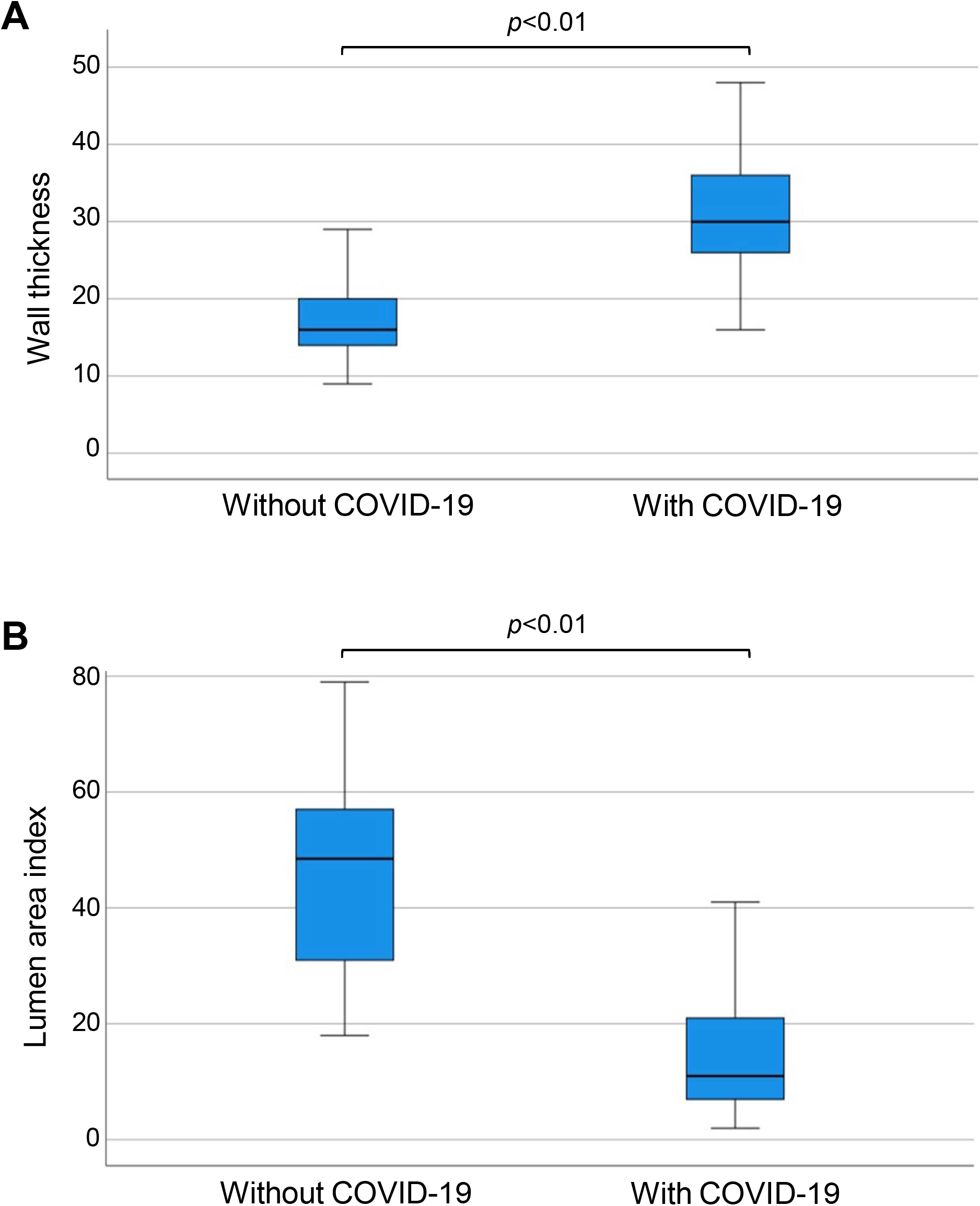
Morphometric analysis of placental arterial wall thickness in women who gave birth without or with COIVD-19. Box plots represent (A) vascular wall thickness and (B) lumen area index values. N = 28 for control and N = 85 for COVID. Mann-Whitney U Test indicated that the two values are significantly different at *p*<0.01.

Masson’s trichrome staining as shown in Fig. 3 shows that women who contracted COVID-19 during pregnancy exhibited severe fibrosis of vascular walls and perivascular space in semi-stem placenta villi.

**Figure 3:**
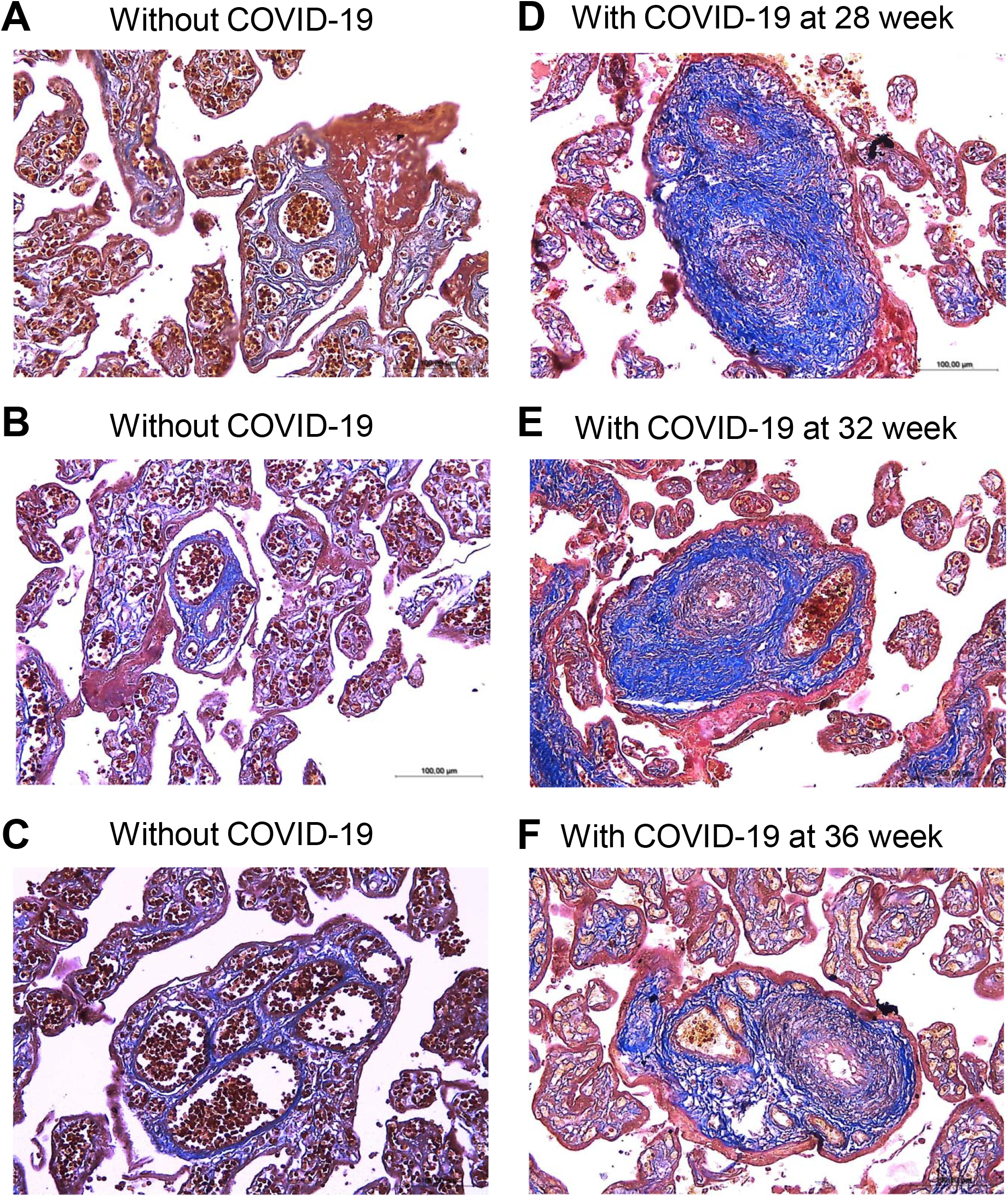
Masson’s trichrome staining. (A-C) Placentas of representative three women who gave birth without having COVID-19 (40 week of gestation). (D-E) Placentas of representative three women who gave birth (40 weeks of gestation) while contracting COVID-19 at 28, 32 or 36 weeks of gestation. Magnification x200.

Immunohistochemistry using a smooth muscle cell marker, α-smooth muscle actin, clearly indicated the dramatic increase in smooth muscle mass in the placental arteries of COVID-19 patients (panels D-F, Fig. 4) compared with controls without COVID-19 (panels A-C, Fig. 4). Given the substantial increase in the mass of smooth muscle, it is likely that the placental arteries of COVID-19 patients underwent smooth muscle cell proliferation.

**Figure 4:**
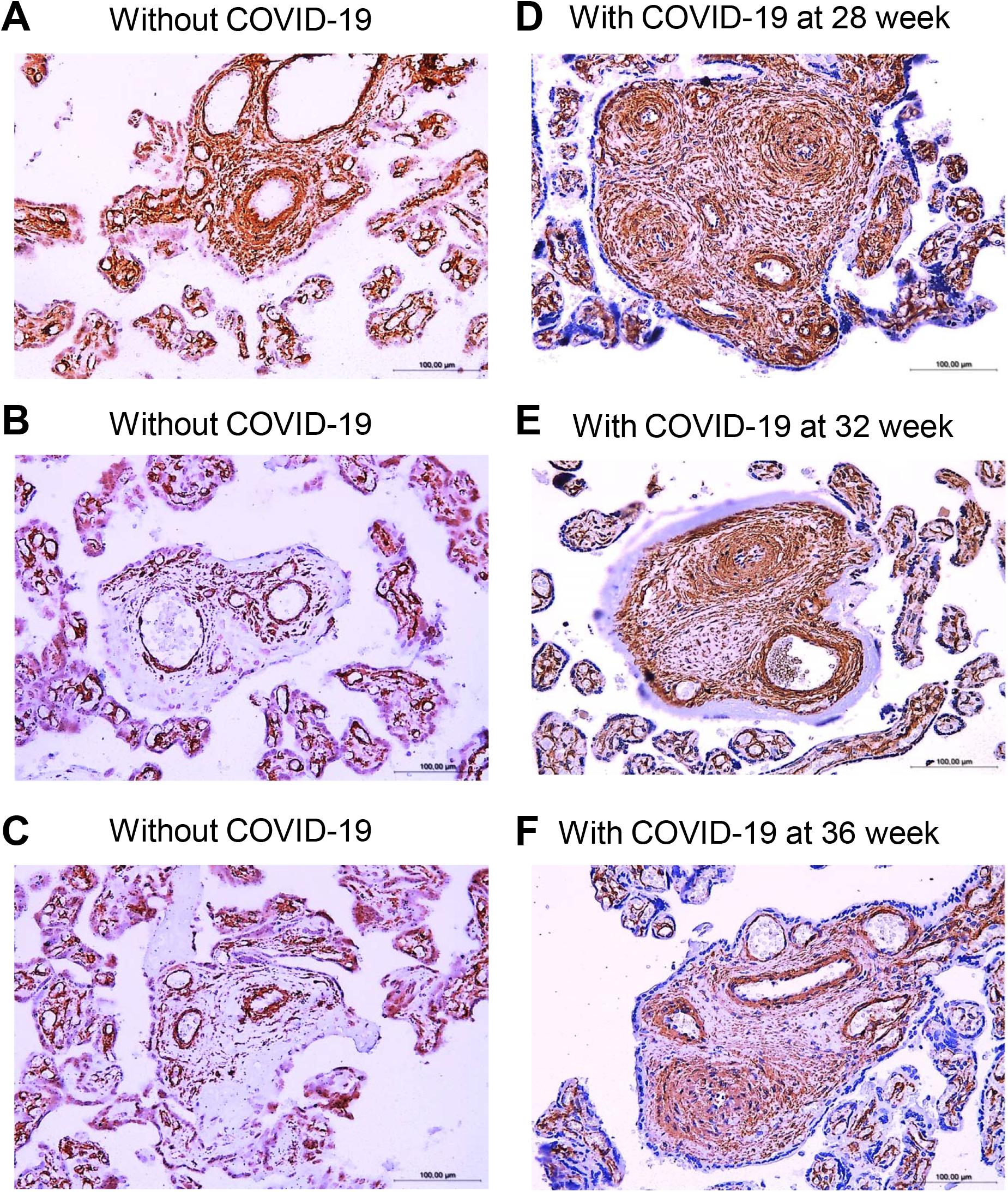
Immunohistochemistry of α-smooth muscle actin in placental arteries of women who gave birth without or with COIVD-19 during pregnancy. (A-C) Immunohistochemistry images of three women who gave birth without contracting COVID-19 (40 weeks of gestation). (D-F) Immunohistochemisry images of three women who gave birth (at 40 weeks of gestation), but contracted COVID-19 at 28, 32, and 36 weeks of pregnancy. Magnification x200.

## Discussion

The major finding of this study is that the arteries of the placentas of women who gave birth to live full-term newborns, but contracted COVID-19 during pregnancy, exhibited severe vascular remodeling.

The narrowing of the placental arterial lumen is expected to alter the blood flow between the mother and the fetus. In the population investigated in this study, despite the occurrence of severe placental vascular remodeling, COVID-19 did not significantly impacted the health of newborn children according to the evaluations of appearance, pulse, grimace, activity, and respiration as quantified by Apgar scores. However, in our participating hospitals in Ukraine, out of 250 births by women who contracted COVID-19 during pregnancy, 13 cases of antenatal fetal death occurred at the rate of 5.2% that is dramatically higher than the average of the general population. Further work should be performed to determine the impact of COVID-19 on pregnancy and the effects of placental vascular remodeling in the well being of the newborns and their development.

Cardiovascular diseases are linked to the development of severe and possibly fatal conditions of COVID-19 [4,9,10]. Thus, the pathology of COVID-19 does not seem to be explained merely by the action of SARS-CoV-2 entering host cells for destruction. Previous studies of the lung vasculature have proposed that SARS-CoV-2 spike protein-mediated cell signaling promotes the hyperplasia and/or hypertrophy of vascular smooth muscle and endothelial cells [14]. More recently, *in vivo* animal studies have confirmed that the spike protein (without the rest of SARS-CoV-2) can adversely influence the vascular system [15]. Further studies investigating the role of the SARS-CoV-2 spike protein in the promotion of placental vascular remodeling would be invaluable to develop effective therapeutic strategies to reduce the mortality and morbidity associated with COVID-19.

In summary, the present study found that placental vascular walls thickened and the lumen narrowed in women who gave live birth but contracted COVID-19 during pregnancy. The smooth muscle proliferation and the collagen fiber deposition appear to play major roles in the development of placental vascular remodeling. Future investigations should aim to determine the role of SARS-CoV-2 spike protein-mediated cell growth signaling in the mechanism of placental vascular remodeling in COVID-19 patients. Elucidating such a mechanism should help provide new therapeutic targets to combat the SARS-CoV-2 infection and COVID-19 and prevent stillbirth and premature birth as well as alternations of child development.

## Data Availability

All data are available from corresponding authors upon request.

## Abbreviations

ACE2: angiotensin-converting enzyme 2
COVID-19: coronavirus disease 2019
H&E: hematoxylin and eosin
SARS-CoV-2: severe acute respiratory syndrome coronavirus 2

